# Hematopoietic fitness of *JAK2^V617F^* Myeloproliferative Neoplasms is linked to clinical outcome

**DOI:** 10.1101/2021.01.28.21250575

**Authors:** Ghaith Abu-Zeinah, Silvana Di Giandomenico, Tatiana Cruz, Elwood Taylor, Ellen K Ritchie, Richard T Silver, Joseph M Scandura

## Abstract

Myeloproliferative Neoplasms (MPN) harbor highly recurrent driver mutations affecting targetable kinases yet treatment options for these phenotypically diverse diseases are limited, and patients experience significant morbidity and shortened survival. The most important disease-related complications—thrombosis, transformation and death—are not used as clinical trial endpoints due to the long follow-up required to assess such disease modifying activity. A reliable monitoring biomarker linking MPN biology with these important clinical outcomes is missing. MPN driver mutation allele frequency (MAF) from whole blood or marrow (WB) does not faithfully predict MPN phenotype, clinical progression or response. This is likely because WB MAF is a composite measure of alleles from a heterogenous and variable mixture of mature leukocytes and, as such, does not report any information about the critical MPN stem and progenitor cells (MPN-SPCs). Driver mutations allow MPN cells to outcompete their normal hematopoietic counterparts and this competitive advantage—increased “fitness”—underlies core biology of MPN pathogenesis. We developed an approach to directly measure MPN fitness from samples. We measured fitness in 115 samples from 84 patients with *JAK2*^*V617F*^ MPNs by quantifying MAF of 11 well-defined and strictly validated hematopoietic stem, progenitor and mature cell populations purified from routinely collected blood and marrow specimens. Unsupervised, hierarchical clustering of MPN fitness revealed 4 major fitness levels: F1, F2, F3, and F4 with significantly different but overlapping clinical features and diagnoses. Notably, these four fitness levels were associated with significantly different event-free survival (EFS): 95% (F1), 81% (F2), 73% (F3), 50% (F4) at 24 months (log-rank p=0.017). In contrast, WB MAF quartile failed to predict EFS. Multivariable models showed that fitness was associated with event risk independent of age, sex, duration of disease, MPN diagnosis and WB MAF. Principal component analysis allowed convenient projection of the 11-component MAF fitness measures to reduce dimensionality and develop a model for relative risk (RR) of event that could be used to assess individual or serial samples. Serial samples with more than a year of follow-up was available for 13 patients. We found that a reduction of this RR score was associated with a therapeutic response (p=0.045). In contrast, increasing RR overtime portended a disease-related event (p=0.045). Changes in WB MAF did not correlate with RR (r^2^=0.022) possibly explaining why WB MAF failed to predict events. These data demonstrate that fitness dynamics from serial blood samples can be used as a monitoring biomarker to assess changes in RR over time. Thus, fitness risk is a promising endpoint alongside corresponding clinical parameters such as blood counts, spleen size and marrow fibrosis grade. Our study offers a feasible approach to monitor the MPN biology central to disease progression and can be used in clinical trials to efficiently identify disease-modifying, potentially life-prolonging treatments.

---

Classical Myeloproliferative Neoplasms (MPN) are chronic, phenotypically diverse diseases^1^ associated with significant morbidity, shortened survival^2^, and limited treatment options^3^. Development of life-prolonging, potentially curative drugs for MPNs has been more challenging than expected given that the vast majority of patients harbor a highly recurrent driver mutation that activates receptor tyrosine kinase signaling. Clinical trials have avoided important clinical endpoints—thrombosis, progression and mortality—because these events can take years or decades to develop. There are presently no reliable monitoring biomarkers to predict these outcomes or assess disease-modifying activity of study agents.

MPNs are initiated by clonal acquisition of a driver mutation by a hematopoietic stem cell (HSC)^4,5^ and progress because the malignant MPN stem and progenitor cells (MPN-SPCs) can outcompete their normal hematopoietic counterparts^5^. Over 90% of MPNs have a driver mutation in just one of three genes (JAK2, CALR and MPL)^6^ and driver mutation allele frequency (MAF) in whole blood or marrow (WB) can provide a crude estimate of tumor burden. However, WB MAF does not reliably distinguish clinical phenotypes or predict outcomes^7–9^. Driver mutations augment JAK/STAT pathway signaling in MPN-SPCs, lineage restricted precursor cells, and mature effector cells; thereby increasing their “fitness” to outcompete normal hematopoietic cells and drive MPN phenotypes. The fitness advantage conferred by MPN driver mutations appears to be lineage and differentiation-stage specific, thereby uncoupling WB VAF from MPN-SPC fitness. Indeed, the competitive advantage of terminally differentiating myeloid MPN cells can be disproportionately higher than that of MPN-SPCs^10–12^ due to augmented cytokine signaling in rapidly proliferating precursor cells^5,13^. Factors linked to MPN phenotypes and prognosis^6,10,11,14–18^ such as driver mutation copy number ^10,11,15^, the presence of co-occurring mutations^16^, and the MPN microenvironment^14^ are also likely to modulate MPN fitness.

MPN fitness underlies core biology of MPN pathogenesis. We developed an approach to estimate fitness by sorting routinely collected peripheral blood (PB) and bone marrow (BM) specimens into in 11 well-defined and strictly validated hematopoietic stem, progenitor and mature cell populations (Figure 1A, Supplementary Figure 1 and Supplementary Methods). Driver MAF was quantified by droplet digital PCR (ddPCR) using DNA extracted from purified cells. Between 8/2017 and 1/2021, we measured hematopoietic fitness in 115 specimens collected from 84 patients with a *JAK2*^*V617F*^ MPN (Supplementary Table 1A). Unsupervised, hierarchical clustering of MPN fitness revealed 4 major Fitness Levels: F1, F2, F3, and F4 (Figure 1B). The pattern of *JAK2*^*V617F*^ propagation through hematopoietic lineages differed for the four fitness levels (Supplementary Figure 2). MPN-SPC fitness was lowest in F1 and highest in F4 but the competitive advantage of *JAK2*^*V617F*^ MPN cells was not uniform across groups; granulocytic differentiation increased fitness in F2/F4 but did so only modestly, or not at all, for F1/F3. Thus, variation in MPN hematopoietic fitness was greater, and more patterned, than previously appreciated.

**Figure 1:**
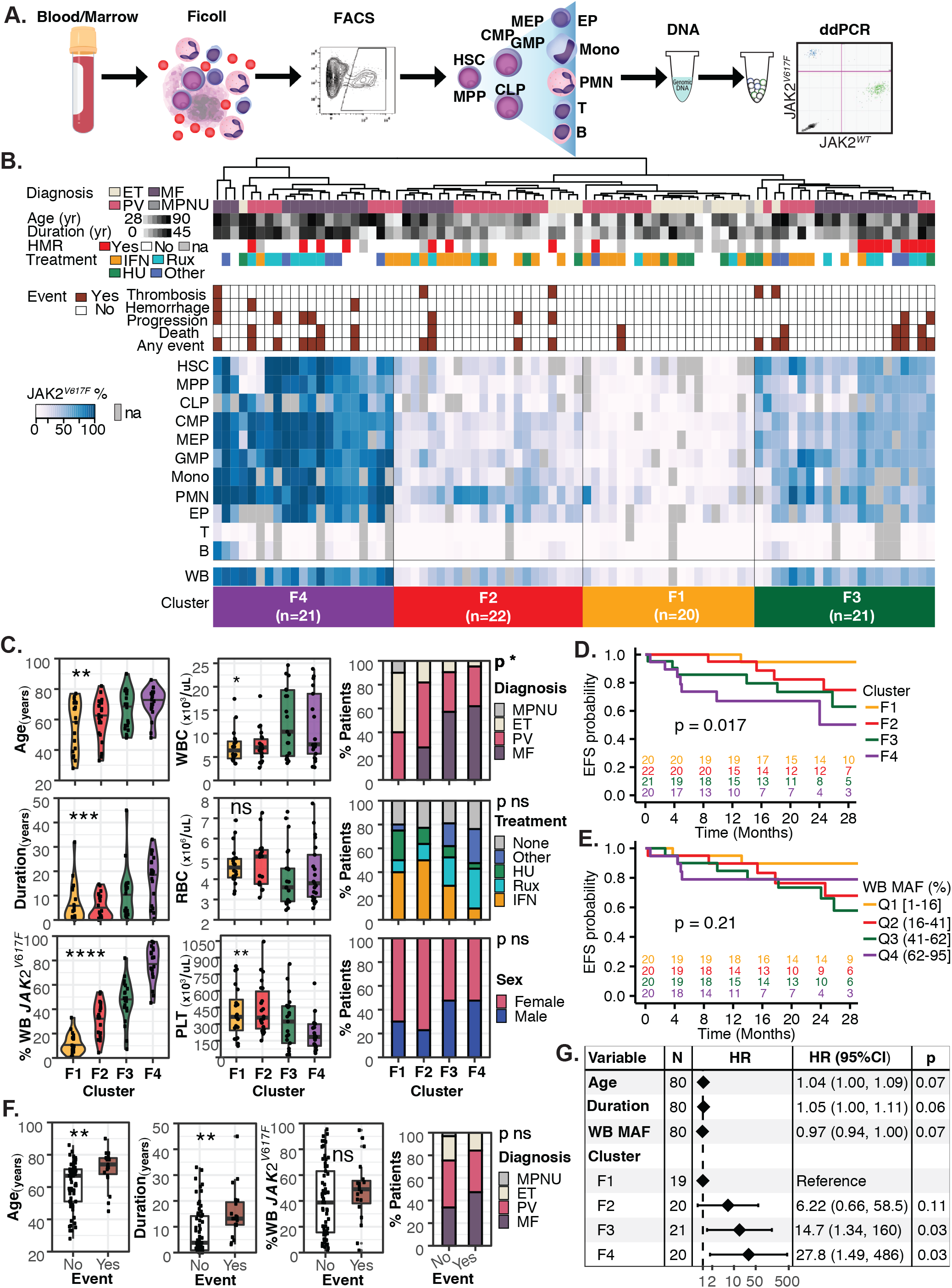
MPN fitness clusters are associated with clinical features and outcome. **A)** Schematic of strategy to purify and assess 11 hematopoietic populations form blood or marrow: hematopoietic stem cell (HSC), multipotent progenitor (MPP), common lymphoid progenitor (CLP), common myeloid progenitor (CMP), megakaryocyte-erythroid progenitor (MEP), granulocyte-macrophage progenitor (GMP), erythroid precursor (EP), monocyte (Mono), neutrophil (PMN), T lymphocyte (T) and B lymphocyte (B). **B**) Heatmap of MPN fitness generated by unsupervised, hierarchical, principal component clustering of 11-population *JAK2*^*V617F*^ MAFs for 84 patients with MPN. Four major fitness clusters (F1, F2, F3, F4) are highlighted with relevant clinical information indicated under the dendogram including diagnosis, age, duration of MPN (duration), high-molecular risk mutation status (HMR), treatment and outcome (event) shown under the dendogram. **C**) Age, duration, WB MAF, white blood cell (WBC) count, platelet (PLT) count, and diagnosis were each significantly different across clusters whereas red blood cell (RBC) counts, treatment distribution, and sex were not. **D**) Event-free survival (EFS) was significantly different between fitness clusters (log-rank p=0.017) with the highest EFS in F1 and the lowest in F4. **E**) EFS did not differ between WB MAF quartiles. **F**) Age and duration, but not WB MAF, or diagnosis were significantly different between patients who had an event and patients who did not. **G**) Multivariat**e** Cox model shows that MPN fitness cluster contributed to event risk independent of age, duration, WB MAF. Age, duration and WB MAF were not independently associated with event risk. Statistically significant changes are indicated (* = p<0.05, **= p<0.01, ***= p<0.001, ****= p<0.0001).

**Figure 2:**
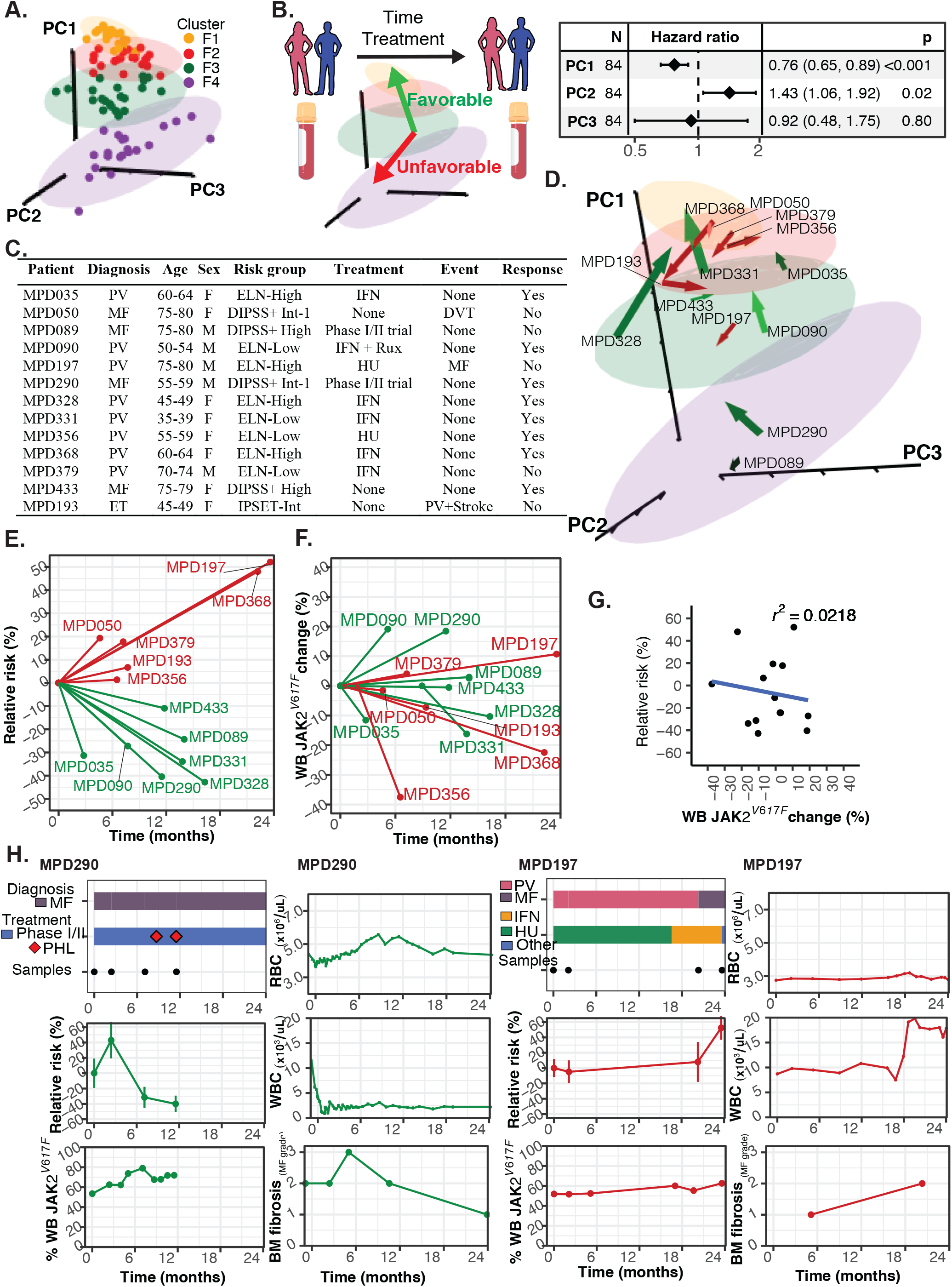
Dynamics of MPN fitness but not WB MAF predict clinical outcomes. **A**) 3-dimensional (3D) projections of the first three principle components (PC) of MPN fitness is shown for the initial sample of 84 patients. Color denotes fitness groups F1 (yellow), F2 (red), F3 (green), and F4 (purple). **B**) Schematic showing favorable or unfavorable MPN fitness dynamics in the 3D PC space and the Cox proportional-hazards model linking PCs to relative risk (RR) of an event, thereby linking fitness dynamics to clinical outcome. A decrease in PC1 and an increase in PC2 are associated with a significantly higher event risk. **C**) 13 patients with serial samples are listed by patient ID; diagnosis; age; sex; risk group (European LeukemiaNet (ELN) for PV, Dynamic International Prognostic Scoring System plus (DIPSS-plus) for MF, and International Prognostic Score for Thrombosis (IPSET) in ET); Treatment such as interferon-alpha (IFN), ruxolitinib (Rux), hydroxyurea (HU); Event such as deep vein thrombosis (DVT) of progression from ET to PV (PV) and PV to MF (MF). **D**) MPN fitness dynamics of the 13 patients in 3D space shown as PC vectors in favorable direction (green arrows) and unfavorable direction (red arrows). **E**) Event RR decreased in seven patients, three of whom started treatment with IFN for PV (MPD035, MPD331, MPD328), one started IFN+Rux for PV (MPD090), and two enrolled on a clinical trial for MF (MPD290, MPD089). Event RR increased in 6 patients including one who later developed MF (MPD197), one who developed PV and had a stroke (MPD193) and one who developed a DVT (MPD050). **F**) Change in WB MAF of the same 13 patients. **G**) WB MAF changes did not correlate with RR changes. **I**) Examples from patients MPD290 and MPD197 showing diagnosis, treatment, relative risk, WB MAF, RBC, WBC, and BM fibrosis over time. Changes in RR reflect changes in clinical and histologic parameters more closely than WB MAF.

As expected, clinical variables were linked to MPN fitness (Figure 1C, and Supplementary Table 1). Patients with more fit MPNs (F3/F4) tended to be older and were more likely to have abnormal blood counts at the time of sampling. These patients also tended to have had an MPN for a longer duration and a higher WB MAF. The proportion of patients with essential thrombocythemia (ET), polycythemia vera (PV), myelofibrosis (MF) and MPN unclassifiable (MPN-U) varied somewhat among groups, with more indolent diseases predominating F1/F2. Although MPN treatment did not strictly correlate with fitness, patients treated with an interferon were more common in F1/F2 (p=0.002, Fisher test). Importantly, event-free survival (EFS) differed significantly across the four fitness levels (p=0.017) with F1 having the longest EFS (95% at 24 months) and F4 the shortest (50% at 24 months) (Figure 1D). In contrast, WB MAF quartile was not associated with EFS (Figure 1E). Patients experiencing an adverse event were older and had MPNs for a longer duration (Figure 1F). Neither WB MAF nor MPN diagnosis differed between those experiencing an event and those that did not. MPN fitness was associated with event risk independent of age, sex, diagnosis, MPN duration, and WB MAF in multivariable time-to-event analysis (Figure 1I and Supplementary Figure 3), suggesting that MPN fitness carries prognostic information.

The event risk associated with a specimen’s fitness was often higher, or lower, than the patient’s clinical diagnosis indicated. We found several patients in unexpectedly high-risk fitness groups who subsequently progressed, or experienced other adverse events after sample collection. In contrast, the subset with MPN fitness lower than expected was enriched for patients receiving interferon; potentially reflecting disease-modifying therapeutic effects in these clinically stable patients^19^. To reduce data dimensionality, we performed principle component analysis (PCA) of the 11-population MAF measures. The first three component vectors (PC1-3) explained 87% of the variance between samples (Supplementary Figure 4). Sample locations in the 3D space of the first three principle components clustered according to MPN clonal fitness groups (Figure 2A). Cox proportional-hazards modeling linked both PC1 and PC2 to event risk (Figure 2B). Whereas PC1 was moved by fitness (allelic burden) in MPN-SPCs and myeloid lineages, PC2 was driven by lymphoid fitness (Supplemental Figure 4). These results potentially explain why WB MAF—predominated by alleles from neutrophils—was not associated with event risk (Figure 1F-G and Supplementary Figure 5).

We monitored MPN fitness longitudinally in patients to establish whether fitness dynamics correlate with clinical response or event occurrence. We used the PCA Cox model to predict relative risk (RR) so that PCA position could be converted to RR on a linear scale. Serial samples were available from 13 patients: 1 with ET, 8 with PV and 4 with MF (Figure 2C). Although 3D PCA projections of serial samples were largely static for patients with stable disease, we identified several patients with unexpectedly large deviations in this space suggesting a change in MPN fitness (Figure 2D). Normalizing risk to the first fitness measurement, we calculated the change in apparent risk from the earliest to the last specimen (Figure 2F). Whereas decreased RR >5% was associated with therapeutic response (p=0.045, Fisher Test), increased RR >5% portended an MPN-related event (3 patients; 2 transformed and 2 had thrombosis, p=0.045, Fisher Test). Interestingly, changes in WB MAF did not correlate with those of RR (r^2^=0.022, Figure 2F-G) possibly explaining why WB MAF failed to predict events. These data demonstrate that serial blood samples can be used to monitor RR over time alongside corresponding clinical parameters such as blood counts, spleen size and marrow fibrosis grade (Figure 2H and Supplemental Figures 6-8).

Biomarkers are needed to sensitively and robustly monitor risk of clinically-important MPN outcomes such as progression, thrombosis and death. Without validated monitoring biomarkers, we are left with crude clinical measures that fall short as treatment decision-making tools. Our study offers a feasible approach to monitor the MPN biology central to disease progression. This approach can be used in clinical trials to efficiently identify therapies with the potential to modify disease outcomes important to patients and clinicians^20^. We found that peripheral blood mononuclear cell (PBMC) populations yielded fitness measures indistinguishable from those including PMNs (supplementary Figure 9) thereby vastly simplifying future use of cryopreserved specimens. MPN fitness measurement also promises to improve prediction of MPN morbidity and progression by reporting individualized disease risk. Prospective studies with long-term outcomes are needed to realize this potential as a biomarker for disease-modification and progression in MPNs. Mechanistic studies to decipher the complex biology of clonal fitness are required to identify the most promising therapeutic approaches to reduce the competitive advantage of MPN-SPCs and improve outcomes for MPN patients.

## Data Availability

No external data sets

## Author contributions

GAZ – designed the study, performed the experiments, examined and consented patients, collected data, analyzed the data and wrote the manuscript.

SDG – performed the experiments and reviewed the manuscript ETIII – collected data.

EKR and RTS – examined and consented patients and reviewed the manuscript

JMS – conceived and designed the study, examined and consented patients, analyzed the data and wrote the manuscript

## Acknowledgements

We acknowledge Mr. Spencer Krichevsky, Ms. Niamh Savage, Ms. Gabriela Hoberman, Ms. Claudia Sosner, and Ms. Diana Jaber from the Joint Clinical Trials Office of Weill Cornell for their exceptional efforts in consenting patients and collecting research specimens.

