## Supplementary Methods for "Hematopoietic fitness of *JAK2^V617F^* Myeloproliferative Neoplasms is linked to clinical outcome"

Ethics Statement

The study was approved by the Institutional Review Board of Weill Cornell Medicine (WCM) and informed consent was obtained in writing from all participating patients in accordance with the Declaration of Helsinki. All samples were de-identified at the site of collection by clinical research staff and given a unique donor ID. The donor ID was linked to the medical record number hosted on a Research Electronic Data Capture (REDCap, 10.0.28-Vanderbilt University) secure database at WCM. The donor ID is only available to the research team and is not publicly available or known to patients.

Sample collection

Peripheral blood (PB) and bone marrow (BM) aspirate specimens were prospectively collected at the Richard T. Silver Myeloproliferative Neoplasms Center of WCM between August 2017 and January 2021 from patients with Ph-neg MPNs. MPN hematopoietic fitness was measured in 115 samples collected from 84 patients with *JAK2^V617F^* MPN (Supplementary Table1a).

Density partitioning and CD34+ immunomagnetic selection

PB samples were centrifuged to separate plasma from the cellular compartment whereas BM samples were filtered through a 70 µm cell strainer (352350, Corning). The cellular compartment was resuspended 1:1 with PBS buffer containing 0.2% BSA and 2mM EDTA and carefully layered over Ficoll (Ficoll-Paque PLUS, Cytiva, formerly GE Healthcare Life Sciences) for density gradient centrifugation (20-minute, 400g spin with no brakes). The Ficoll buoyant mononucleated (MNC) and the Ficoll dense polymorphonucleated (PMN) cell layers were collected. The PMN layer was treated with 10x red cell lysis buffer (BioLegend). The MNCs were stained with CD34 MicroBeads (Miltenyi Biotec) and then immunomagnetic selection for CD34+ cells was performed using LS columns (Miltenyi Biotec) and MidiMACS separator (Miltenyi Biotec). Both CD34+ and CD34-depleted fractions were collected.

Immunostaining and FACS

All fluorochrome conjugated antibodies were from Biolegend except where specified. The PMN layer was stained with CD45, CD15 (BD Biosciences), and CD16. CD34- cell fraction was divided into two; one was stained with CD45, CD3, CD19 (Beckman Coulter), CD11b (BD Biosciences), and CD14; and the other was stained with CD123 (Thermo Fisher Scientific), CD36 (BD Biosciences), CD71 (BD Biosciences), and biotinylated anti-human lineage (Lin) antibodies (CD3, CD14, CD15, CD19, CD235a), followed by secondary staining with BUV395-Streptavidin (BD Biosciences). The CD34+ fraction was stained with CD45, CD34, CD38, CD10, CD45RA, CD90, CD123, and biotinylated anti-human lineage antibodies (CD3, CD14, CD15, CD19, CD235a), followed by secondary staining with BUV395-Streptavidin (BD Biosciences).

Final purification of specimens was performed by multiparameter FACS (FACSAria II, BD Biosciences) to isolate CD45^+^CD15^+^CD16^+^ polymorphonucleated cells (PMNs), CD45^+^CD14^+^CD11b^+^ monocytes, CD45^+^CD3^+^ T cells, CD45^+^CD19^+^ B cells, and CD45^+^Lin^-^CD123^-^CD36^+^CD71^+ 1^ erythroblasts from the CD34- fraction. The CD34+ cells were FACS sorted into 6 well-defined HSPC populations^2,3^ using a 6-way sorter (Influx, BD Biosciences) into CD45^dim^Lin^-^CD34^+^CD38^-^ CD45RA^-^CD90^+^ (HSC), CD45^dim^Lin^-^CD34^+^CD38^-^ CD45RA^-^CD90^-^ (MPP), CD45^dim^Lin^-^CD34^+^CD38^+^CD10^+^ (CLP), CD45^dim^Lin^-^CD34^+^CD38^+^CD45RA^-^CD123^+^ (CMP), CD45^dim^Lin^-^CD34^+^CD38^+^CD45RA^-^CD123^-^ (MEP), CD45^dim^Lin^-^CD34^+^CD38^+^CD45RA^+^CD123^+^ (GMP). Viable cells were sorted based on DAPI staining (un-fixed cells) or zombie (PFA fixed cells). The FACS gating strategy is shown in Supplementary Figure 1 (FCS Express 7, De Novo Software).

DNA extraction, droplet digital PCR (ddPCR) and quantification of mutant allele frequency (MAF)

DNA was extracted from the FACS sorted populations and whole blood or marrow (WB) using the QIAamp DNA Blood Mini kit in accordance with the manufacturer's protocol (51106, Qiagen). For small samples, DNA QIAamp DNA Micro Kit was used (56304, Qiagen). Up to 50 ng of genomic DNA (gDNA) in 8µL of nuclease-free water was loaded per well on a 96-well plate with 11µL of Supermix for probes (1863024, Bio-Rad) and 1 µL of *JAK2^V617F^/ JAK2^WT^* primer-probe assay (10049551 or dHsaMDV2010061, Bio-Rad) for a total volume of 20 µL/well. The Bio-Rad QX200 ddPCR system with automated droplet generator was used to generate ddPCR droplets and analysis. Thermocycling was performed as recommended (Bio-Rad). The number of droplets positive for *JAK2^V617F^* and *JAK2^WT^* alleles and total droplets were determined using the Bio-Rad QX200 ddPCR system analyzer and Bio-Rad Quantasoft software. The MAF and 95% confidence intervals were estimated using Poisson statistics (R, rateratio package). Samples that yielded < 10 total positive droplets were excluded from the final analysis because of low confidence in the MAF value (Supplementary Figure 9).

Clinical data collection

Data on patient age, sex, diagnosis, symptoms, spleen size, blood counts, bone marrow pathology, molecular pathology, cytogenetics, risk group^4–7^, diagnosis date, treatment and disease-related event dates was collected and hosted on WCM REDCap. Structured query language (SQL) was used for automated data extraction from electronic medical records, facilitating the collection of clinical data. Natural language processing tools (R 4.0.3 and R Studio 1.3.1093 ) were utilized to extract data on spleen size from physical exam documentation and imaging reports; and bone marrow features from the pathology reports^8^. Patients meeting the following WHO 2016 diagnosis criteria^9^ for Ph-neg MPNs were included: essential thrombocythemia (ET); polycythemia vera (PV); or myelofibrosis (MF) for primary myelofibrosis, post-ET and post-PV myelofibrosis; and unclassified MPN (MPNU). High molecular risk (HMR) profile was determined for ET, PV and MF based on mutations in the following sets of genes, respectively: *SF3B1, SRSF2, U2AF1, and/or TP53* ^10^; *SRSF2*^10^; and *ASXL1, SRSF2, IDH1/2, EZH2*^11^. Start and end dates for MPN treatments, and dates of phlebotomy (PHL) were recorded. MPN treatments included interferon alpha (IFN), hydroxyurea (HU), ruxolitinib (Rux) and other (less commonly used drugs such as anagrelide, busulfan and imatinib; combinations such as IFN+Rux; clinical trial treatments such as Rux+navitoclax and CPI-0610). The diagnosis date and dates of disease-related events were recorded through manual review of medical records. Criteria for inclusion were manually reviewed and verified for all patients. Disease-related events were categorized as thrombosis (myocardial infarction, stroke, venous and arterial thromboses), hemorrhage (clinically significant major or non-major bleeding events as defined by the ISTH^12^), disease progression (ET progression to PV defined by WHO criteria^9^, ET or PV progression to MF per IWG-MRT criteria^13^, or any MPN evolving to MDS, accelerated phase MPN or AML per the WHO criteria^9,14^). Time to event was determined from time of sample collection to the event or censoring at last follow-up if no event occurred.

Data analysis

Principal component analysis (PCA) was performed for hierarchical clustering of patients based on 11-dimensional MAF data (dendogram in Figure 1B) and for reduction of dimensions to cluster data points in a 3-dimensional space using the first principal components (PC) 1, 2 and 3 (Figure 2A). WB MAF was excluded from the clustering algorithms. Samples with >5 missing MAFs were excluded. Missing data, as a result of incomplete processing or a yield of < 10 ddPCR droplets, were indicated in the heatmap (Figure 1B) in gray. A fully interpolated matrix for PCA was obtained by imputation of missing values using DINEOF (Data Interpolating Empirical Orthogonal Functions) procedure^15^. The procedure was shown to accurately determine Empirical Orthogonal Functions from data sets with gaps^16^. Major hierarchical clusters were compared by age, sex, diagnosis, treatment, duration of disease, blood counts by descriptive statistics, Fisher’s exact test for distribution of categorical variables and ANOVA for continuous variables. A Wilcoxon signed-rank test was used to compare medians of two groups and a Student’s *t*-test was used to compare means of two groups. The EFS for fitness clusters and WB MAF quantile groups was estimated using Kaplan-Meier (KM) methods and compared between groups using log-rank test. A multivariable Cox proportional-hazards model was used for time-to-event analysis of variables such as fitness cluster and WB MAF.

A multivariable Cox proportional-hazards model was also used for time-to-event analysis of PC1, PC2 and PC3. The model provided an estimate of a patient’s relative risk (RR) of an event at any time based on the PC values derived from the PCA of 11-population MAFs. PCs were calculated for 11-population MAFs of serial samples from individual patients. This enabled serial RR prediction and determination of RR reduction or increase over time. Descriptive statistics were used to visualize a patient’s RR over time alongside the clinical timeline (diagnosis, treatment) and trends in blood counts, spleen size and marrow fibrosis.

All p-values were derived from two-tailed statistical analyses; an alpha level of 0.05 was used. The analysis was performed using RStudio software v 1.1.423 (packages used are listed in reference section).

**R packages references**

- R Core Team (2020). R: A language and environment for statistical computing. R Foundation for Statistical Computing, Vienna, Austria. URL <https://www.R-project.org/>.
- Hadley Wickham, Romain François, Lionel Henry and Kirill Müller (2020). dplyr: A Grammar of Data Manipulation. R package version 1.0.2. <https://CRAN.R-project.org/package=dplyr>
- Zhao, Xiaobei and Sandelin, Albin. GMD: Generalized Minimum Distance of distributions. R package. 2014. [http://CRAN.R-project.org/package=GMD](http://cran.r-project.org/package=GMD)
- Hadley Wickham (2007). Reshaping Data with the reshape Package. Journal of Statistical Software, 21(12), 1-20. URL <http://www.jstatsoft.org/v21/i12/>.
- Daniel Adler, Duncan Murdoch and others (2020). rgl: 3D Visualization Using OpenGL. R package. version 0.103.5. <https://CRAN.R-project.org/package=rgl>
- H. Wickham. ggplot2: Elegant Graphics for Data Analysis. Springer-Verlag New York, 2016.
- Alboukadel Kassambara, Marcin Kosinski and Przemyslaw Biecek (2020). survminer: Drawing Survival Curves using 'ggplot2'. R package version 0.4.8. <https://CRAN.R-project.org/package=survminer>
- Dane R. Van Domelen (2019). tab: Create Summary Tables for Statistical Reports. R package version 4.1.1. <https://CRAN.R-project.org/package=tab>
- Therneau T (2020). _A Package for Survival Analysis in R_. R package version 3.2-7, <URL: <https://CRAN.R-project.org/package=survival>>.
- Terry M. Therneau, Patricia M. Grambsch (2000). _Modeling Survival Data: Extending the Cox Model_. Springer, New York. ISBN 0-387-98784-3.
- Paul Murrell (2014). gridBase: Integration of base and grid graphics. R package version 0.4-7. <https://CRAN.R-project.org/package=gridBase>
- Minato Nakazawa (2019). fmsb: Functions for Medical Statistics Book with some Demographic Data. R package version 0.7.0. <https://CRAN.R-project.org/package=fmsb>
