## Supplemental Tables and Figures for "Hematopoietic fitness of *JAK2^V617F^* Myeloproliferative Neoplasms is linked to clinical outcome"

**Supplementary Table 1: Demographics and clinical features of patients and fitness clusters**

| Variable | Overall (n = 84) | F1 (n = 20) | F2 (n = 22) | F3 (n = 21) | F4 (n = 21) | P |
|---|---|---|---|---|---|---|
| Age, median (range) | 68 (28-90) | 64 (28-77) | 62 (33-82) | 70 (48-90) | 73 (48-86) | 0.010 |
| Female, n (%) | 53 (63) | 14 (70) | 17 (77) | 11 (52) | 11 (52) | 0.22 |
| Diagnosis, n (%) | | | | | | <0.001 |
| ET | 17 (21) | 10 (56) | 4 (18) | 2 (10) | 1 (5) | |
| PV | 34 (41) | 8 (44) | 12 (55) | 7 (33) | 7 (33) | |
| MF | 31 (38) | 0 (0) | 6 (27) | 12 (57) | 13 (62) | |
| Disease duration, median (range) | 6 (0-45) | 4 (0-32) | 3 (0-15) | 11 (0-45) | 19 (0-33) | <0.001 |
| HMR, n evaluable = 63, n(%) | 13 (21) | 1 (7) | 3 (17) | 5 (38) | 4 (22) | 0.23 |
| Treatment, n (%) | | | | | | 0.05 |
| IFN | 27 (32) | 8 (40) | 11 (50) | 6 (29) | 2 (10) | |
| Rux | 17 (20) | 2 (10) | 3 (14) | 5 (24) | 7 (33) | |
| HU | 11 (13) | 5 (25) | 3 (14) | 2 (10) | 1 (5) | |
| Other | 11 (13) | 1 (5) | 0 (0) | 4 (19) | 6 (29) | |
| None | 18 (21) | 4 (20) | 5 (23) | 4 (19) | 5 (24) | |
| RBC, median (range) | 4.2 (2.5-7.7) | 4.6 (3.3-6.9) | 5.1 (3.1-7.3) | 3.6 (2.5-7.3) | 3.8 (2.6-7.7) | 0.07 |
| WBC, median (range) | 8 (3-68) | 6 (4-17) | 7 (3-18) | 10 (4-68) | 8 (3-50) | 0.08 |
| PLT, median (range) | 307 (8-1045) | 366 (102-823) | 358 (151-1045) | 323 (8-843) | 184 (37-668) | 0.003 |
| Samples/patient, median (range) | 1 (1-10) | 1 (1-4) | 2 (1-10) | 2 (1-5) | 1 (1-8) | 0.45 |
| Follow-up duration (months), median (range) | 25 (-0-36) | 28 (5-36) | 28 (0-34) | 23 (0-36) | 13 (-0-34) | 0.02 |

**Supplementary Figure 1: FACS strategy for isolating 11 hematopoietic populations.**
A) CD34+ mononucleated cells FACS sorted into 6 HSPC populations: HSC, MPP, CLP, CMP, MEP, GMP. B) CD34- mononucleated cells FACS sorted into T, B, and Mono. C) CD34- mononucleated cells FACS sorted into EP. D) polymorphonucleated cells sorted into neutrophils. E) morphologic appearance of HSPC and mature cells and functional validation of progenitors by colony forming units: CFU-GEMM from CMP, CFU-GM from GMP and BFU-E from MEP

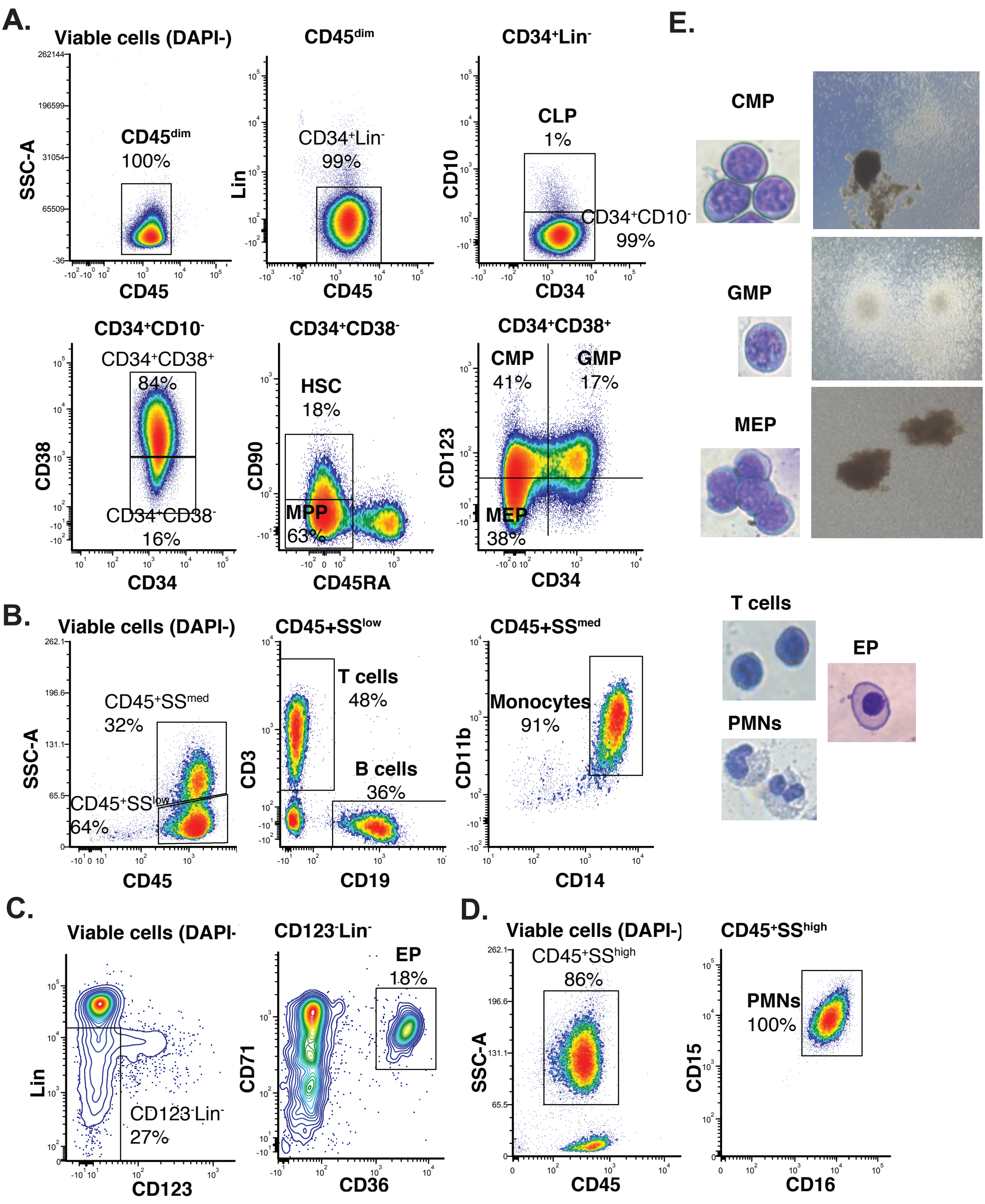

**Supplementary Figure 2:** **Hematopoiesis fitness patterns of *JAK2[V617F]* MPN fitness clusters.**
A) Average fitness pattern in hematopoiesis of clusters F1, F2, F3, and F4 shown as mean patient MAF +/- standard deviation (Stdev) of the 11 populations at different stages and lineages of hematopoietic differentiation. Increasing HSPC MAF seen from F1 to F4. B) Radar plots of F1, F2, F3, F4 showing the difference in mean MAF between HSC+MPP and each of the 9 differentiated progenitors and mature cells with increased myeloid differentiation fitness (blue) most notable in F2/F4 and decreased lymphoid fitness (red) in all clusters. Statistically significant differences are indicated (* = p<0.05, **= p<0.01, ***= p<0.001).

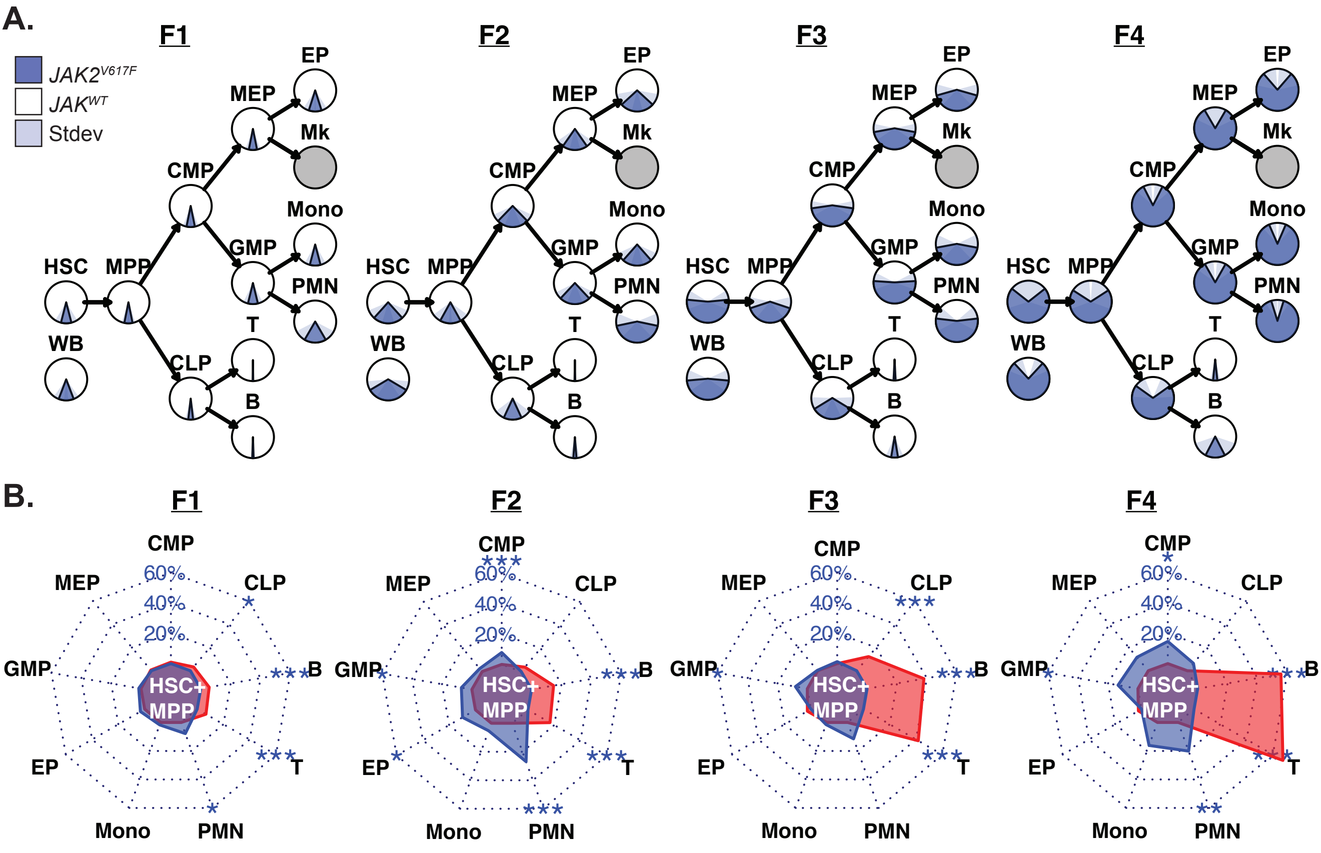

**<u>Supplementary Figure 3:</u> Multivariable analysis of time to event**

Results show statistically significant association of fitness clusters with risk of event, independent of age, sex, diagnosis, duration, and WB MAF.

| Variable | N | Hazard ratio | | p |
|---|---|---|---|---|
| **Age** | 80 | | 1.04 (1.00, 1.09) | 0.06 |
| **Sex** | | | | |
| F | 50 | | Reference | |
| M | 30 | | 1.39 (0.45, 4.30) | 0.57 |
| **Diagnosis** | | | | |
| ET | 16 | | Reference | |
| PV | 31 | | 0.71 (0.16, 3.14) | 0.65 |
| MF | 31 | | 0.56 (0.11, 2.95) | 0.49 |
| MPNU | 2 | | 0.00 (0.00, Inf) | 1.00 |
| **Duration** | 80 | | 1.04 (0.98, 1.10) | 0.19 |
| **WB MAF** | 80 | | 0.97 (0.93, 1.00) | 0.07 |
| **Cluster** | | | | |
| F1 | 19 | | Reference | |
| F2 | 20 | | 8.89 (0.70, 112.59) | 0.09 |
| F3 | 21 | | 22.33 (1.43, 348.26) | 0.03 |
| F4 | 20 | | 54.88 (1.41, 2143.23) | 0.03 |

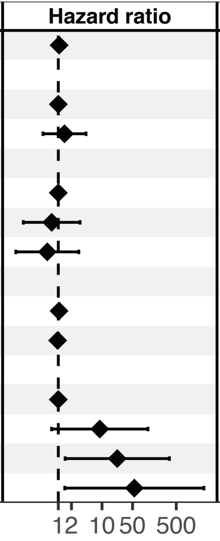

**Supplementary Figure 4: Correlation of 11-population MAF with principal components 1-3.** Principal component (PC) 1 explains 71% of variance and is inversely correlated with all 11 population MAF, but primarily HSPC and myeloid lineages whereas PC2 explains 11% and is directly correlated with T and B cell MAFs. PC3 is inversely correlated with HSC+MPP, not correlated with progenitors (CLP, CMP, MEP, GMP) and loosely with mature myeloid cell MAF (EP, Mono, PMN).

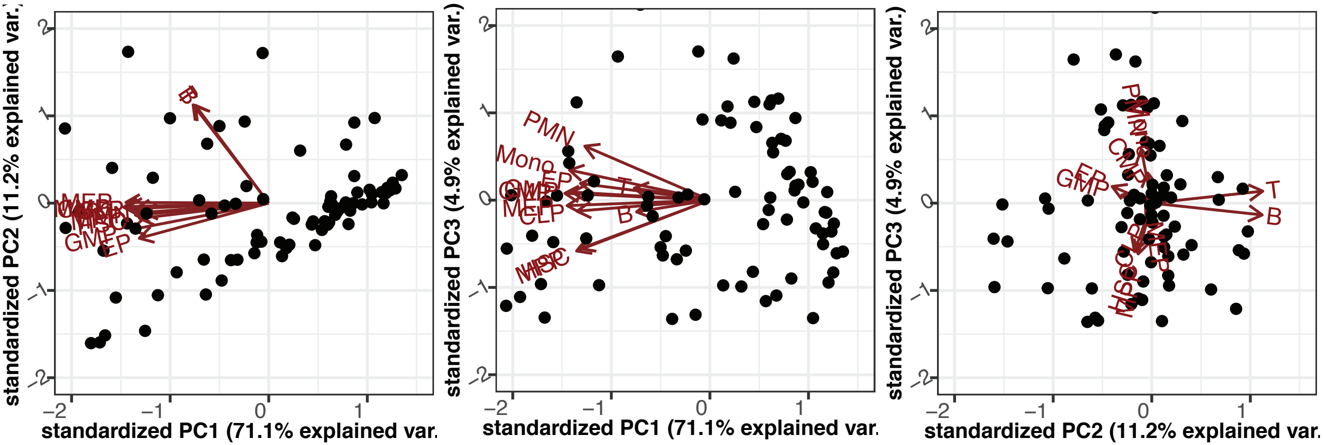

**Supplementary Figure 5: Correlation of individual population MAF with event and correlation of 10-population MNC, 6-population HSPC and 5-population mature cells with 11-population MAF.** A) Patients who had an event were more likely to have higher HSC, MPP, CLP, CMP, MEP, Mono and T cell MAFs, but not GMP, PMN, EP , B cell or WB MAFs. B) All except WB and EP MAFs were associated with increased risk of event in univariable time-to-event analysis. C) Schematic posing the question of whether HSPC, MNC or Mature cell fitness alone correlate with11-population MAF. E) near-perfect correlation in predicted risk of event from MNC fitness with 11-population fitness and no correlation between HSPC or mature cell fitness with 11-population fitness

**A.**

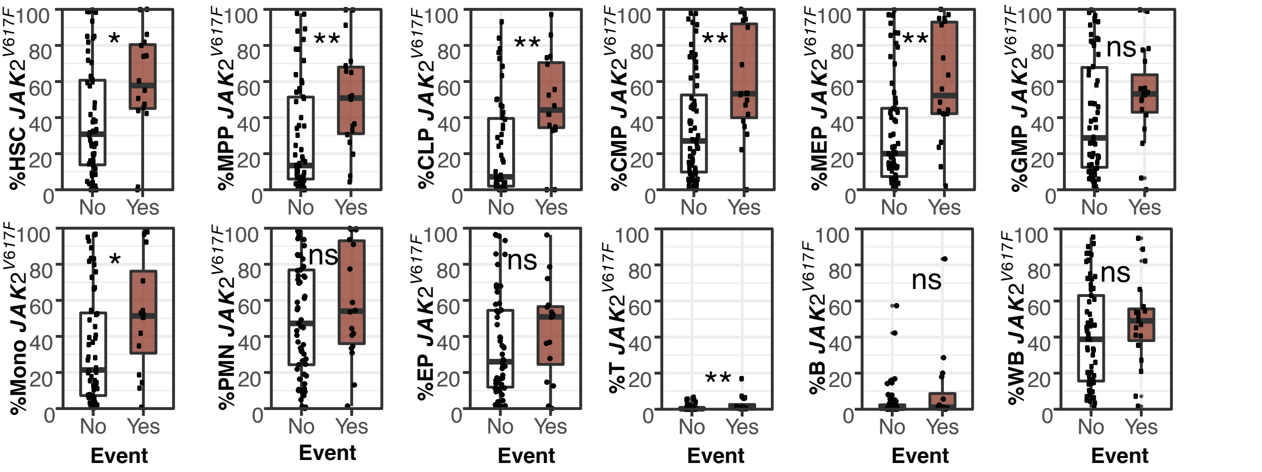


**B.**

| Pop | JAK2$^{V617F}$% Mean (SD) | Univariable HR (95% CI, p-value) |
|---|---|---|
| HSC | 43.3 (31.8) | 1.02 (1.00-1.04, p=0.015) |
| MPP | 33.3 (30.5) | 1.02 (1.01-1.04, p=0.002) |
| CLP | 28.9 (29.0) | 1.03 (1.01-1.05, p=0.001) |
| CMP | 40.6 (31.0) | 1.03 (1.01-1.04, p<0.001) |
| MEP | 37.7 (31.9) | 1.03 (1.01-1.04, p<0.001) |
| GMP | 42.9 (31.4) | 1.01 (1.00-1.03, p=0.036) |
| Mono | 37.9 (32.2) | 1.02 (1.01-1.04, p=0.008) |
| PMN | 52.0 (31.4) | 1.02 (1.00-1.03, p=0.046) |
| EP | 36.5 (28.7) | 1.01 (1.00-1.03, p=0.087) |
| T | 1.3 (2.6) | 1.37 (1.16-1.62, p<0.001) |
| B | 5.5 (13.3) | 1.04 (1.01-1.07, p=0.009) |
| WB | 42.0 (27.3) | 1.02 (1.00-1.03, p=0.066) |

**C.**

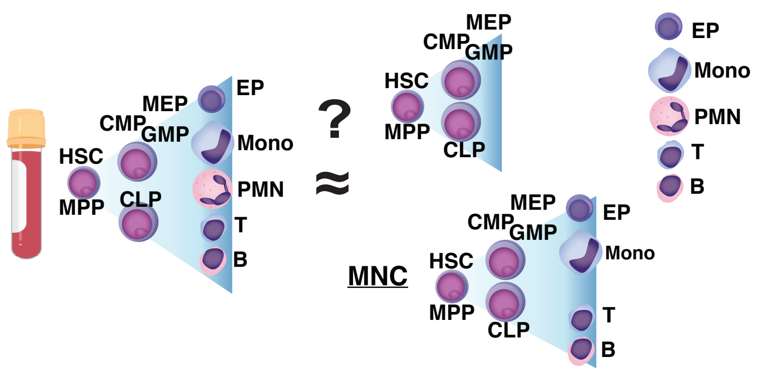


**D.**

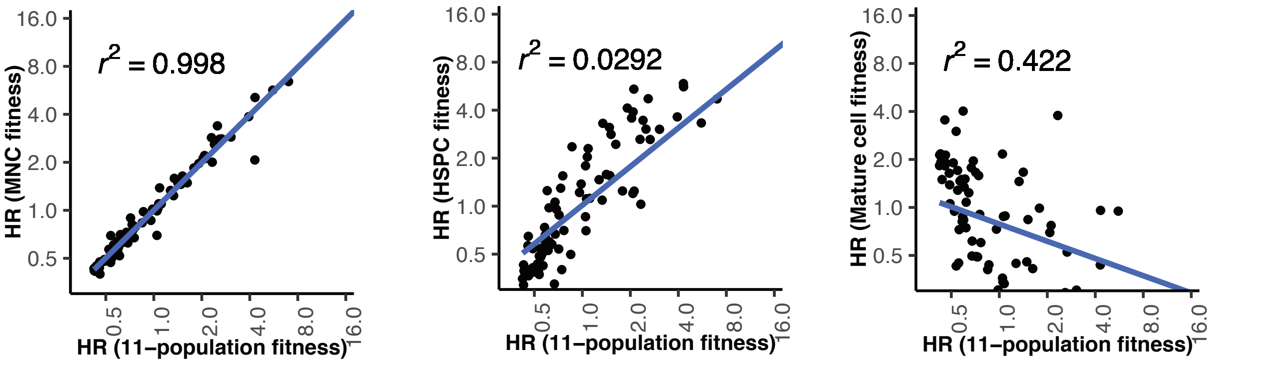

**<u>Supplementary Figure 6:</u> fitness predicted relative risk and clinical variables over time for patients MPD197, 290, 098, and 090.** A) Diagnosis and treatment timeline. B) 3D PCA vectors from serial sample fitness measurement. C) Predicted relative risk of event alongside WB MAF, WBC, RBC, PLT, and BM fibrosis grade.

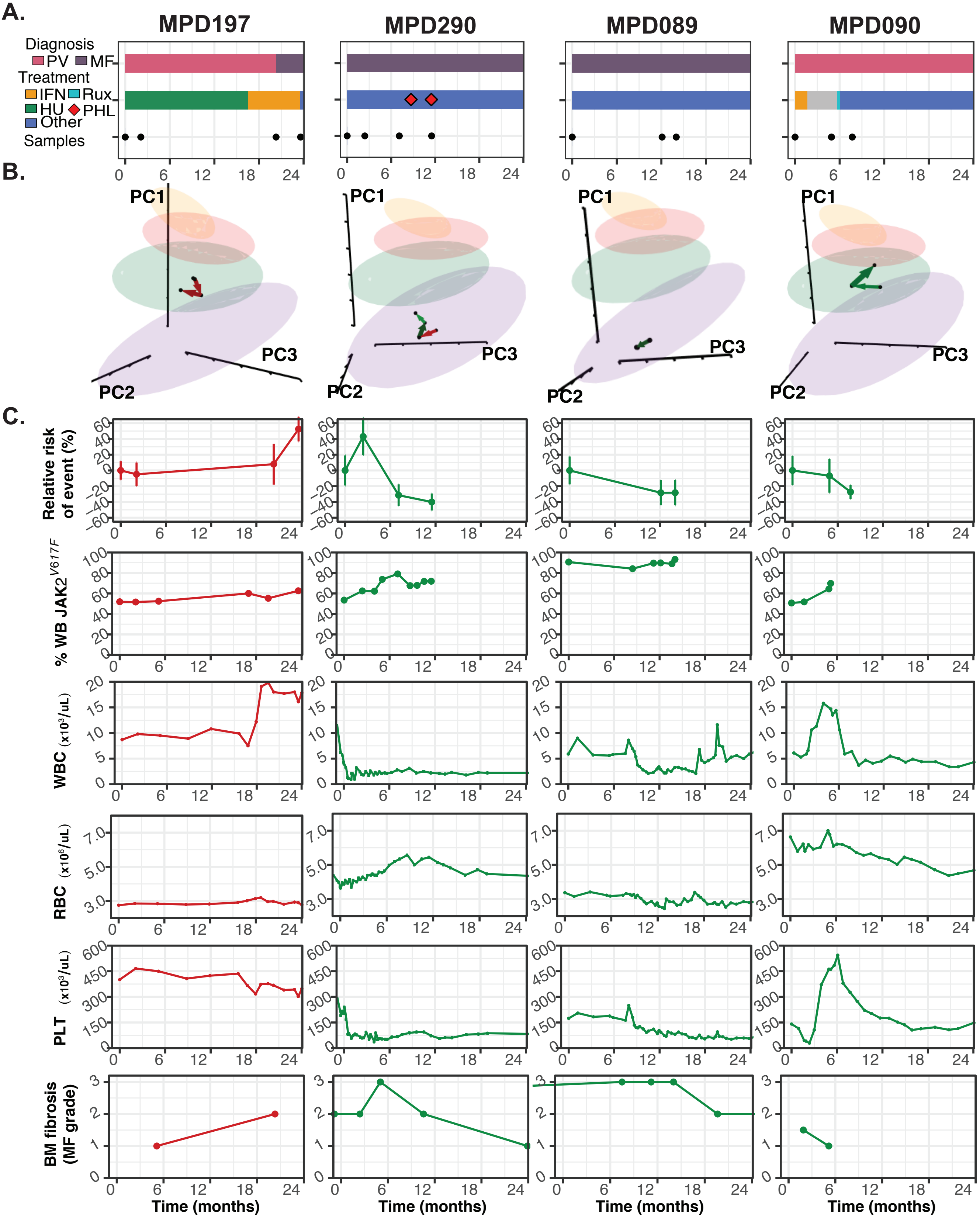

**Supplementary Figure 7: fitness predicted relative risk and clinical variables over time for patients MPD035, 328, 331, and 379.** A) Diagnosis and treatment timeline. B) 3D PCA vectors from serial sample fitness measurement. C) Predicted relative risk of event alongside WB MAF, WBC, RBC, PLT, and BM fibrosis grade.

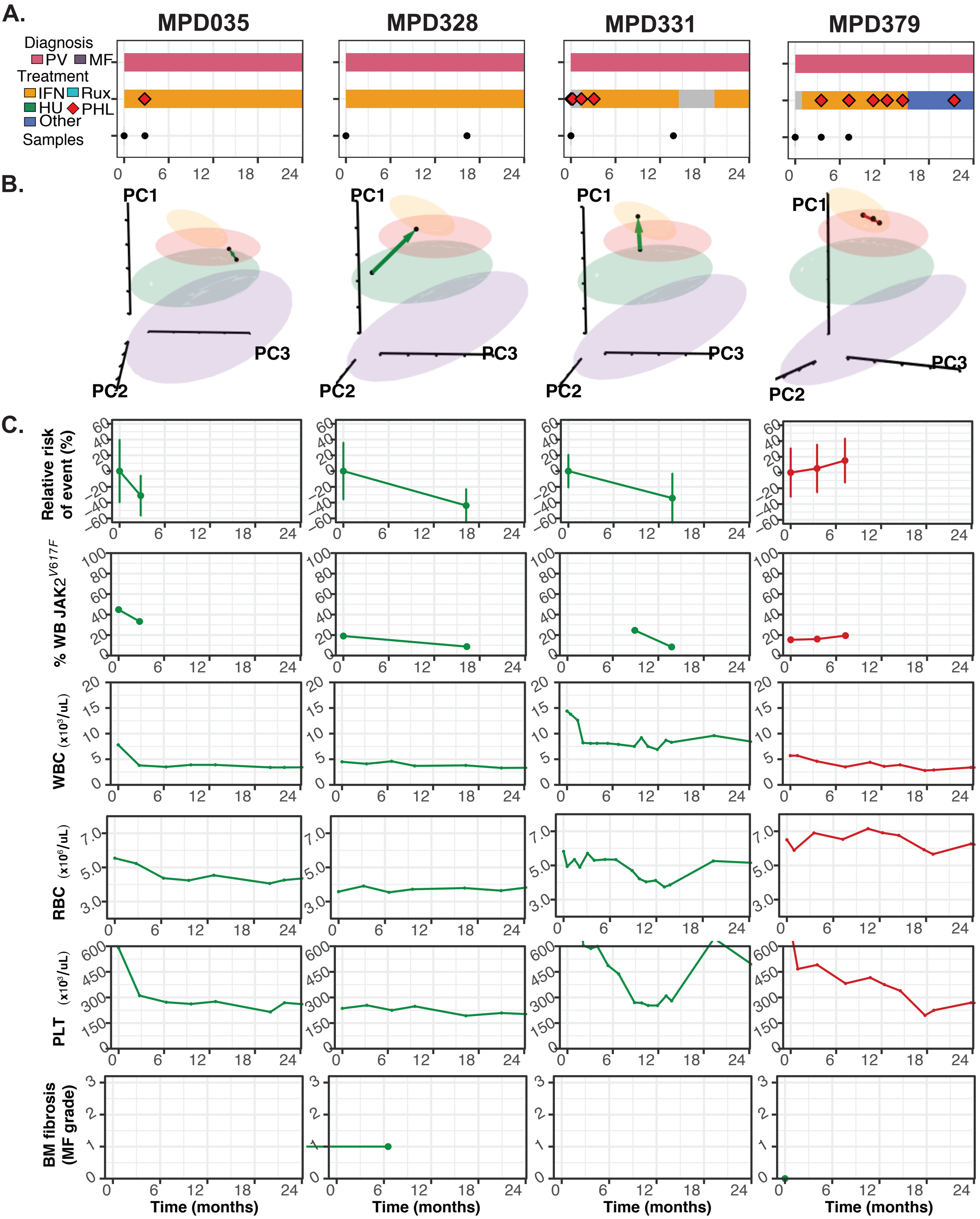

**Supplementary Figure 8: fitness predicted relative risk and clinical variables over time for patients MPD050, 433, 356, and 368.** A) Diagnosis and treatment timeline. B) 3D PCA vectors from serial sample fitness measurement. C) Predicted relative risk of event alongside WB MAF, WBC, RBC, PLT, and BM fibrosis grade.

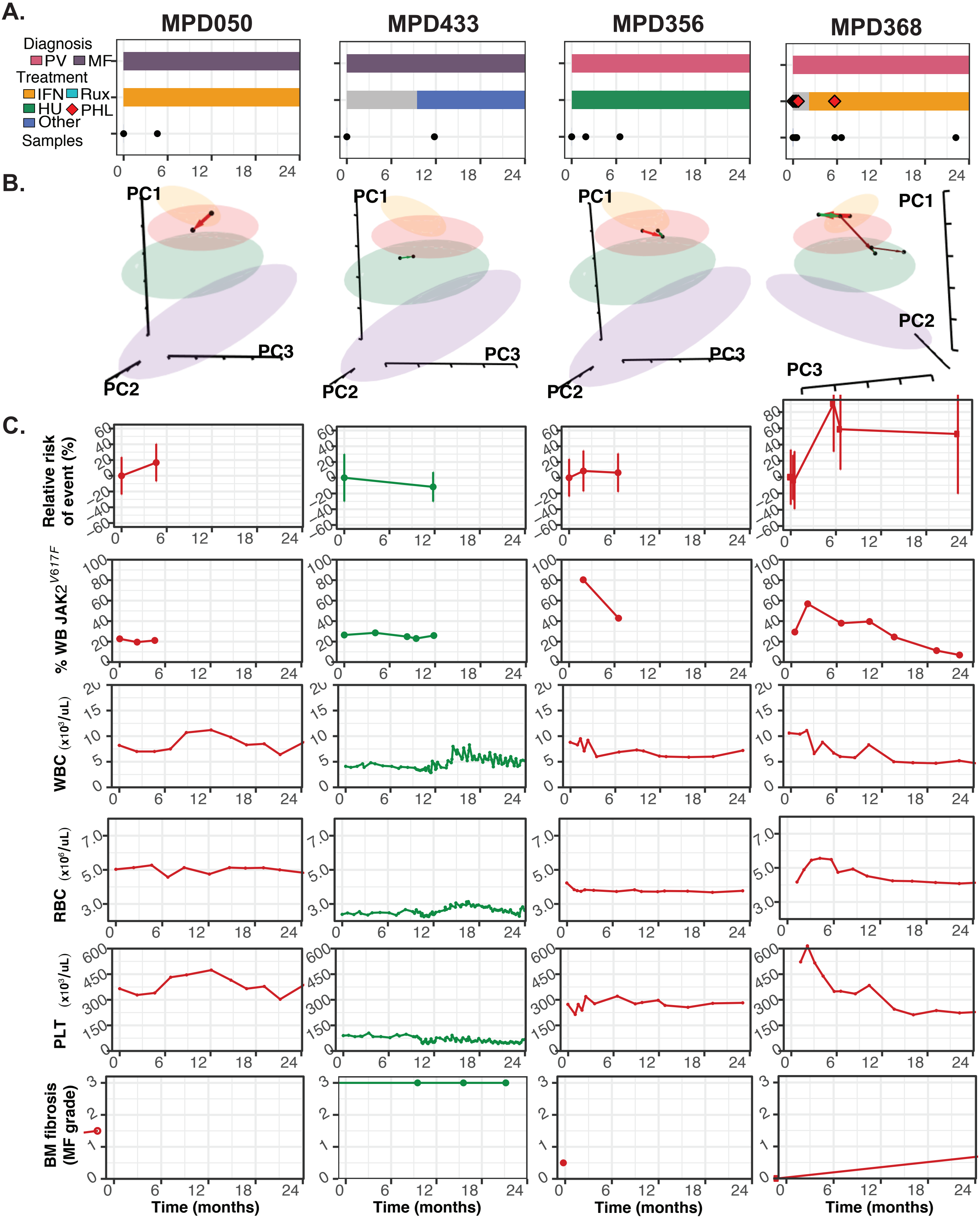
